# Factors associated with the decay of anti-SARS-CoV-2 neutralizing antibodies among recipients of an adenoviral vector-based AZD1222 and a whole-virion inactivated (BBV152) vaccine in Chennai, India: a prospective, longitudinal, cohort study

**DOI:** 10.1101/2022.02.26.22271097

**Authors:** Sivaprakasam T Selvavinayagam, Yean Kong Yong, Hong Yien Tan, Zhong-Ying Zhang, Gurunathan Subramanian, Manivannan Rajeshkumar, Kalaivani Vasudevan, Priyanka Jayapal, Krishnasamy Narayanasamy, Dinesh Ramesh, Sampath Palani, Marie Larsson, Shankar Esaki M, Sivadoss Raju

**Affiliations:** State Public Health Laboratory, Directorate of Public Health and Preventive Medicine, DMS Campus, Teynampet 600 018, Chennai, Tamil Nadu, India; Laboratory Centre, Xiamen University Malaysia, 43900 Sepang, Selangor, Malaysia; School of Traditional Chinese Medicine, Xiamen University Malaysia, 43900 Sepang, Selangor, Malaysia; Chemical Engineering, Xiamen University Malaysia, 43900 Sepang, Selangor, Malaysia; Government Corona Hospital, Guindy, Chennai 600 032, India; Department of Biomedicine and Clinical Sciences, Linkoping University, 58185 Linköping, Sweden; Infection Biology, Department of Life Sciences, Central University of Tamil Nadu, Thiruvarur 610 005, India

**Keywords:** AZD1222, BBV152, COVID-19, IgG decay, SARS-CoV-2, Vaccination

## Abstract

**Background:** The magnitude of protection conferred after recovery from COVID-19 or by vaccine administration, and the duration of protective immunity developed, remains ambiguous.

**Methods:** We investigated the factors associated with antibody decay in 519 individuals who received treatment for COVID-19-related illness or received COVID-19 vaccination with two commercial vaccines, viz., an adenoviral vector-based (AZD1222) and a whole-virion-based inactivated (BBV152) vaccine in Chennai, India from March 2021. Blood samples collected during regular follow-up post-infection/vaccination andwere examined for anti-SARS-CoV-2 IgG by a commercial automated chemiluminescent immunoassay (CLIA).

**Results:** Age and underlying comorbidities were the two variables that were independently associated with the development of breakthrough infection. Individuals who were >60 years of age with underlying comorbid conditions had a ∼15 times and ∼10 times greater risk for developing a breakthrough infection and hospitalization, respectively. The time elapsed since the first booster dose was associated with attrition in anti-SARS-CoV-2 IgG, where each month passed was associated with an ebb in the neutralizing antibody levels by a coefficient of -6 units.

**Conclusions:** Our findings advocate that the elderly with underlying comorbidities require a second booster dose with AZD1222 and BBV152.

## Introduction

The COVID-19 pandemic has caused an unprecedented global crisis, and having lasted for more than two years, has resulted in over 340 million COVID-19 cases, claiming >5.5 million deaths by January 2022, causing huge levels of economic damage and yet there are no signs of waning [1, 2]. While antiviral agents against SARS-CoV-2, the virus causing COVID-19, are yet to become available, vaccines and public health interventions remain the most promising approach against this global peril. Although anti-SARS-CoV-2 vaccines are successful in reducing the rates of infection, and the level of neutralizing antibodies has correlated with protection against SARS-CoV-2 reinfection [3] as well as severe COVID-19,4-6 breakthrough infections do continue to recur. Importantly, it still remains a conundrum, how long this vaccine-induced acquired immunity would last in an individual. Several studies have investigated the dynamics and the duration of protection of neutralizing antibodies developed following the onset of natural SARS-CoV-2 infection or vaccination [7-11]. Notwithstanding, the results thus far remain inconsistent, with some reporting rapid waning of the antibodies months after exposure to the virus or following the administration of a vaccine [7-11], while others reporting the prolonged presence of neutralizing antibodies [12-14]. This discrepancy partly stems from differences in the demography of patients as well as the different vaccines available globally, and hence the magnitude of protection conferred after recovery from a natural infection or by vaccines, and the duration of protective immunity developed post-vaccination, remains ambiguous. Here, we investigated the factors associated with antibody decay following both natural infection as well as vaccination with two commercial vaccines, viz., a viral vector-based AZD1222 and an inactivated BBV152 vaccine.

## Patients and Methods

### Patients

We conducted a population-based study among Chennai’s adult who received treatment for COVID-19-related illness or received COVID-19 vaccination at the State Public Health laboratory, Directorate of Public Health and Preventive Medicine, Chennai, India, and the Government Corona Hospital, Chennai, India from March 2021 until December 2011. The inclusion criteria were that the participants needed to be >18 years, and there were no exclusion criteria. The medical records of the participants were reviewed and data such as patient demography, comorbidities, history of SARS-CoV-2 infection, date and type of vaccine received were recorded. Blood samples were collected during regular follow up post infection/vaccination, and tested for their levels of anti-SARS-CoV-2 IgG. The study was approved by the Human Ethics Committee of the Madras Medical College (EC No. 03092021).

### Anti-SARS-CoV-2 IgG Chemiluminescent assay

Blood collected were tested for their levels of anti-SARS-CoV-2 IgG by VITROS anti-SARS-CoV-2 IgG assay, a commercial automated chemiluminescent immunoassay (CLIA), according to manufacturer’s instructions using a VITROS Anti-SARS-CoV-2 IgG Calibrator on the VITROS ECi/ECiQ/3600 Immunodiagnostic Systems and the VITROS 5600/XT 7600 Integrated Systems. The assay targeted to the spike protein S1 antigen and the cut-off (minimum detection limit) was ≥1.00.

### Statistical analyses

The primary analysis was to compare patients with natural infection, those who received AZD1222 and BBV152. Comparison of categorical variables was tested using the chi-square test, whereas continuous variables (e.g., age) were compared using the unpaired t-test. Potential risk factors for breakthrough infection and hospitalization such as demographics between those who had natural infection, or received AZD1222 and BBV152 vaccination were evaluated by simple and adjusted binary logistic regression. The odds ratio and 95% confidence interval (CI) were estimated. The predictive power of age in predicting breakthrough infection and hospitalization were examined using receiver operating characteristic (ROC) analysis. The decay of decay of anti-SARS-CoV-2 IgG levels was assessed using simple and adjusted linear logistic regression. Statistical analyses were performed using Prism, version 5.02 (GraphPad Software, San Diego, CA). Binary regression was performed using SPSS, version 20 (IBM, Armonk, NY), Two-tailed P <0.05 was considered as statistical significance for all test performed and P value <0.05, <0.01, <0.001, <0.0001 were marked as *, **, *** and ****, respectively.

## Results

### Patients and specimens

We investigated 519 individuals who received treatment for COVID-19-related illness or received COVID-19 vaccination at the State Public Health laboratory, Directorate of Public Health and Preventive Medicine, Chennai, India, and the Government Corona Hospital, Chennai, India from March 2021. The inclusion criteria were that the participants needed to be >18 years, and there were no exclusion criteria. The medical records of the participants were reviewed and data such as patient demography, comorbidities, history of SARS-CoV-2 infection, date and type of vaccine received were recorded. Blood samples were collected during regular follow up post infection/vaccination, and tested for their levels of anti-SARS-CoV-2 IgG. Blood collected were tested for their levels of anti-SARS-CoV-2 IgG by a commercial automated chemiluminescent immunoassay (CLIA). The study was approved by the Human Ethics Committee of the Madras Medical College (EC No. 03092021).

Five hundred and nineteen individuals recruited into the study were separated into two groups, i.e., “unvaccinated” (n=52) versus “vaccinated” (n=467). The “unvaccinated” group was further bifurcated into two sub-groups with those who did not have a history of natural infection of SARS-CoV-2 (n=25) and those who had a natural COVID-19 infection (n=27).

In India, two vaccines were initially approved for administration to the public, one the adenoviral vector vaccine AZD1222 (ChAdOx1) manufactured by the Serum Institute of India, Pune, and the other, a whole-virion inactivated BBV152 vaccine developed by Bharat Biotech International Limited in collaboration with the Indian Council of Medical Research [15]. The vaccinated group was also divided into two groups i.e., participants who received the AZD1222 (n=259) and those who received the BBV152 (n=208) vaccines. Of note, a small fraction of vaccinees had developed a natural infection prior to receiving vaccination (n=85); whilst another portion of the participants had developed a breakthrough infection ∼14 days post-vaccination (n=149). The colored boxes in **Figure 1** represent the three main study groups in our investigation. The median age of the cohort was 34 years with an interquartile range (IQR) of 26–52, with 47.8% male participants. Of note, 74 (14.3%) of the participants had some form of underlying comorbid conditions such as hypertension (n=37), diabetes mellitus (n=24), and heart disease (n=4). Of all the participants, 176 (33.9%) were infected by SARS-CoV-2 (**Supplementary Table 1**). There was a significantly higher number of individuals with COVID-19 among the non-vaccinees (51.9%) as compared to vaccinees (AZD1222=31.7% and BBV152=32%) (P=0.016), indicating that vaccination confers protective immunity among vaccine recipients. There was no significant difference between the percentage of individuals developing breakthrough infection with both the vaccines, indicating that the protective efficacy of both the vaccines are similar and the onset of a breakthrough infection appears to be owing to inadequate cross-neutralizing potential conferred by the vaccine to the circulating virus variants.

**Figure 1:**
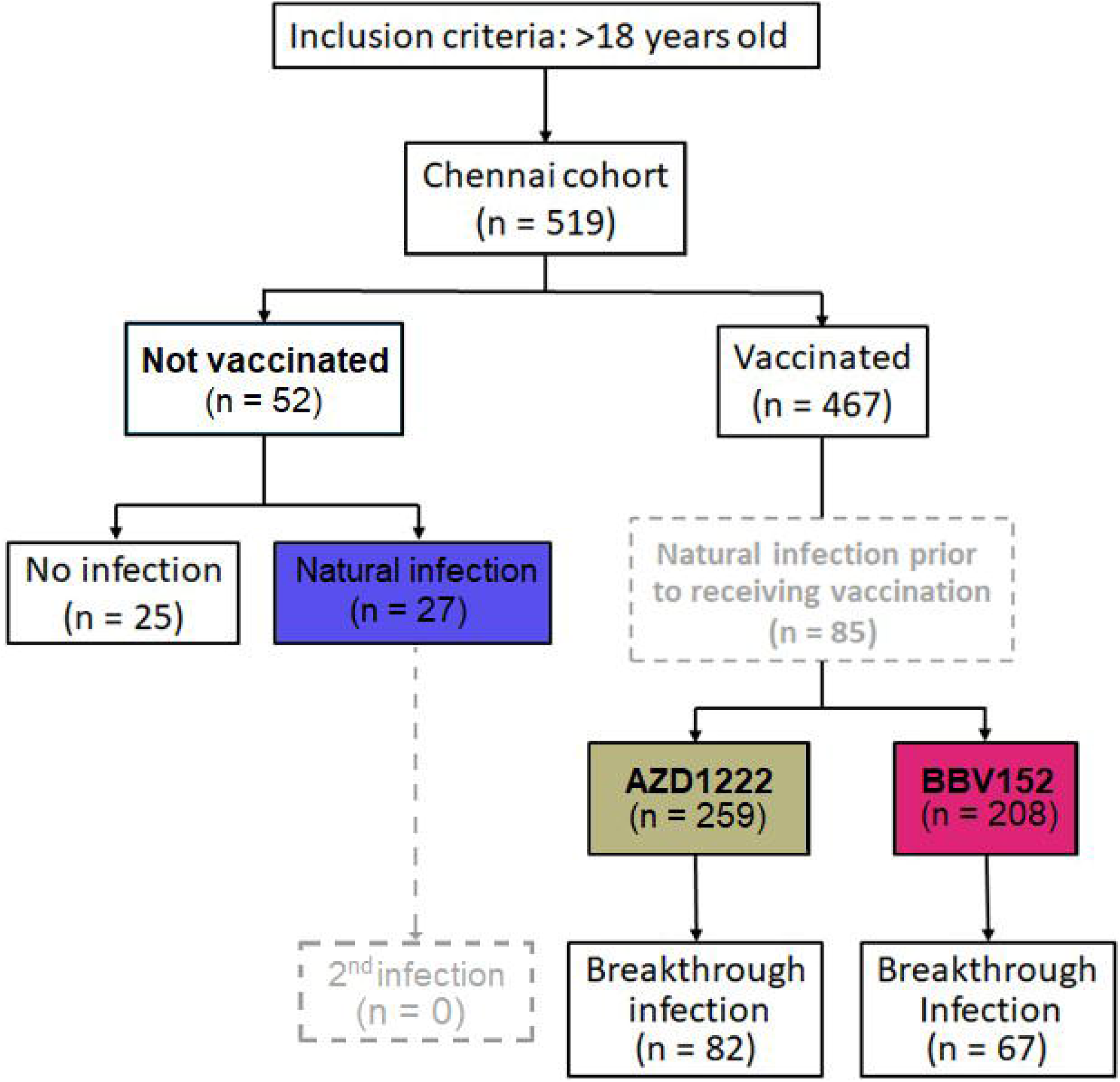
Flow diagram of 519 participants recruited into the Chennai Cohort from March 2021. Based on the sequence of natural infection and type of vaccine administered, the cohort is divided into 4 groups viz., i) No vaccination, no natural infection, ii) no vaccination, with natural infection, iii) vaccination with AZD1222 and iv) with BBV152. Of note, a small fraction of vaccine recipients had a documented history of natural infection prior to receipt of the corresponding vaccine (n=85); whilst another group of participants developed a breakthrough infection 14 days post-vaccination (n=149). The colored boxes, present three main patient groups in the investigation.

Since a minor proportion of individuals develop breakthrough infections despite administration with two doses of the vaccine, we sought to investigate the factors that determine/predict the onset of a breakthrough infection. The association between breakthrough infection and other demographic parameters such as age, gender, occupation as healthcare workers, type of vaccine received and comorbidities were first assessed univariately using a binary regression model. Variables with P value <0.05 will then be included in the multivariate analysis. Variables with P value <0.05 were considered as independent predictors. Our multivariate analysis showed that age and underlying comorbidities were the two variables that were independently associated with development of a breakthrough infection. We also found that every increase of age by 5 years was associated with an increased risk of developing a breakthrough infection by 1.23 unit (95% CI=1.11–1.38; P<0.0001). An existing comorbid condition was associated with an increased risk of contracting a breakthrough infection by 2.07 units (95% CI=1.11–3.89; P<0.023) (**Figure 2A**). We also found that age and comorbidity were independently associated with hospitalization (**Supplementary Table 2**).

**Figure 2:**
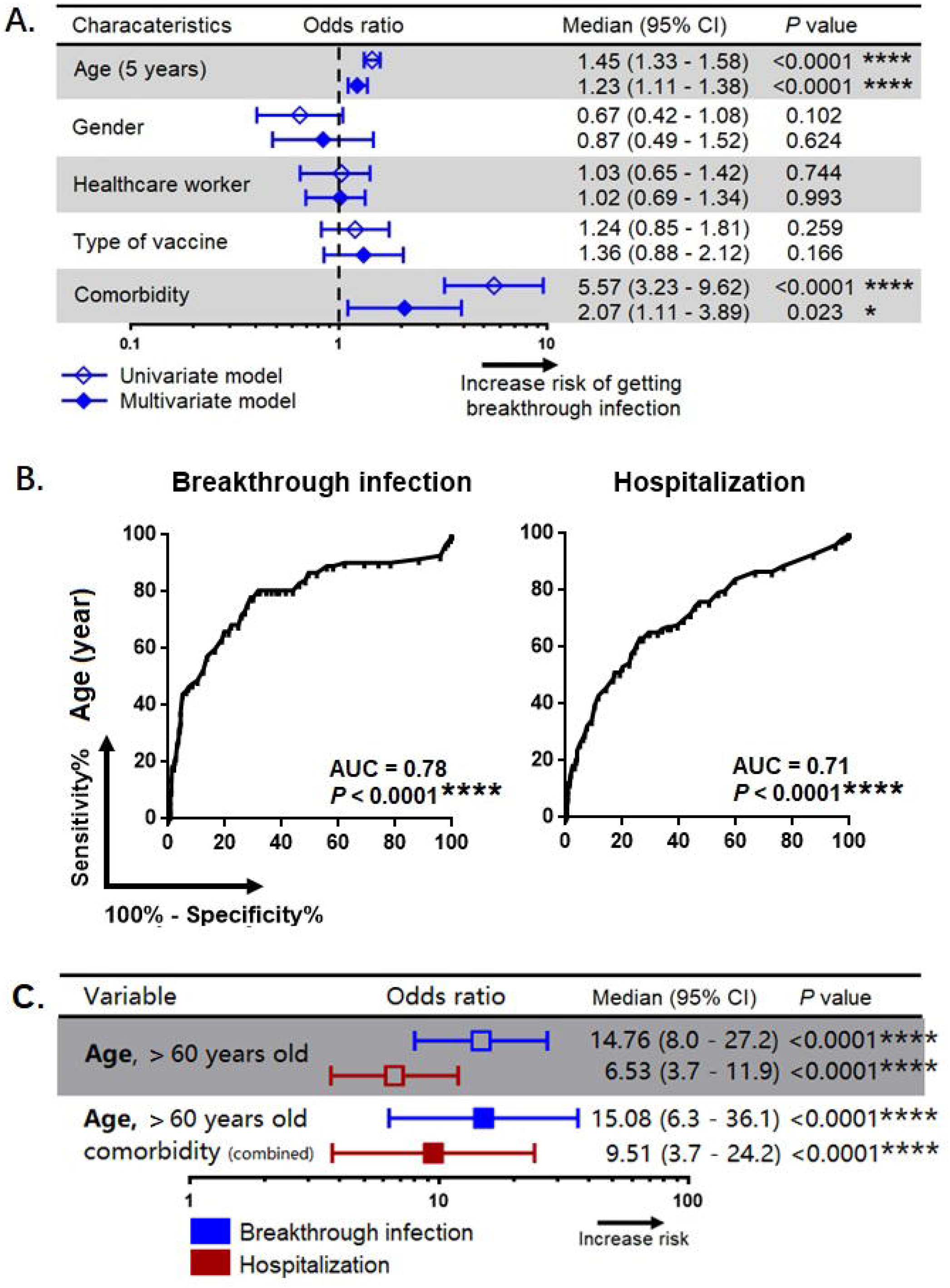
Association of patients’ characteristics with risk for development of breakthrough infection and hospitalization. **A)** A simple and adjusted binary regression models assessing the factors that associated with breakthrough infection. Odds ratios for values below or above threshold levels were displayed in a forest plot; median and 95% CI were calculated. **B)** Receiver operating characteristics analysis for prediction of breakthrough infection and hospitalization. **C)** Association of age and comorbidity with the risk of breakthrough infection and hospitalization. CI, confidence interval. *, **, ***, *** represent *P* <0.05, <0.01, <0.001, <0.0001 respectively.

The receiver operating characteristic curve (ROC) analysis revealed that age was a strong predictor of development of a breakthrough infection (area under curve, AUC=0.78; P<0.0001) as well as hospitalization (AUC=0.71; P<0.0001) (**Figure 2B**), and the cut-off age was determined as 60 years. Our binary regression analysis showed that participants who were >60 years of age and with underlying comorbid conditions had a ∼15 times and ∼10 times greater risk for developing a breakthrough infection and hospitalization, respectively (**Figure 2C**).

Given that the titer of antibodies will decay gradually with time, next we investigated the decay of neutralizing IgG in individuals who experienced a natural infection, and having vaccinated with AZD1222 and BBV152. In this analysis, we only included those who had a natural infection (without vaccination, n=11), and those who had been vaccinated (without natural infection prior to vaccination or a breakthrough infection, n=291). Using a linear regression model, we first studied the decay of anti-SARS-CoV-2 IgG levels across the two vaccines in comparison with natural infection in controlling the days since recovering from a natural infection or after administration with the first booster dose of the respective vaccine.

The univariate analysis showed that the time elapsed since the first booster dose of the vaccine was associated with the reduction in the levels of anti-SARS-CoV-2 IgG, where each month passed was associated with reduction in the neutralizing antibody levels by a coefficient of -6 unit (95% CI=-9.88–-2.1; P=0.003) (**Table 1**). In the adjusted model, we found that participants who were >60 years of age had an accelerated decay rate, where each day of lapse was associated with a decrease of IgG by a coefficient of 23 units (95% CI=-46.69–-0.05; P=0.047). However, such decay of IgG was only observed among participants who received AZD1222, but not among those who received BBV152 and those who had a history of recovery from a natural SARS-CoV-2 infection (**Figure 3A**). Using binary regression, we assessed the time elapsed since vaccination and their association with development of a breakthrough infection and hospitalization, controlling for age (>60 years). Here, we showed that time elapsed since vaccination was an independent predictor of development of a breakthrough infection and hospitalization in those who had received the AZD1222 vaccine. As the level of anti-SARS-CoV IgG gradually decays with time, our regression model showed that each month of lapse was associated with increased risk of contracting a breakthrough infection and hospitalization by 0.85 (95% CI=0.72–1.01; P=0.048) and 0.85 (95% CI=0.73–0.98; P=0.041), respectively (**Figure 3B**).

**Table 1:**
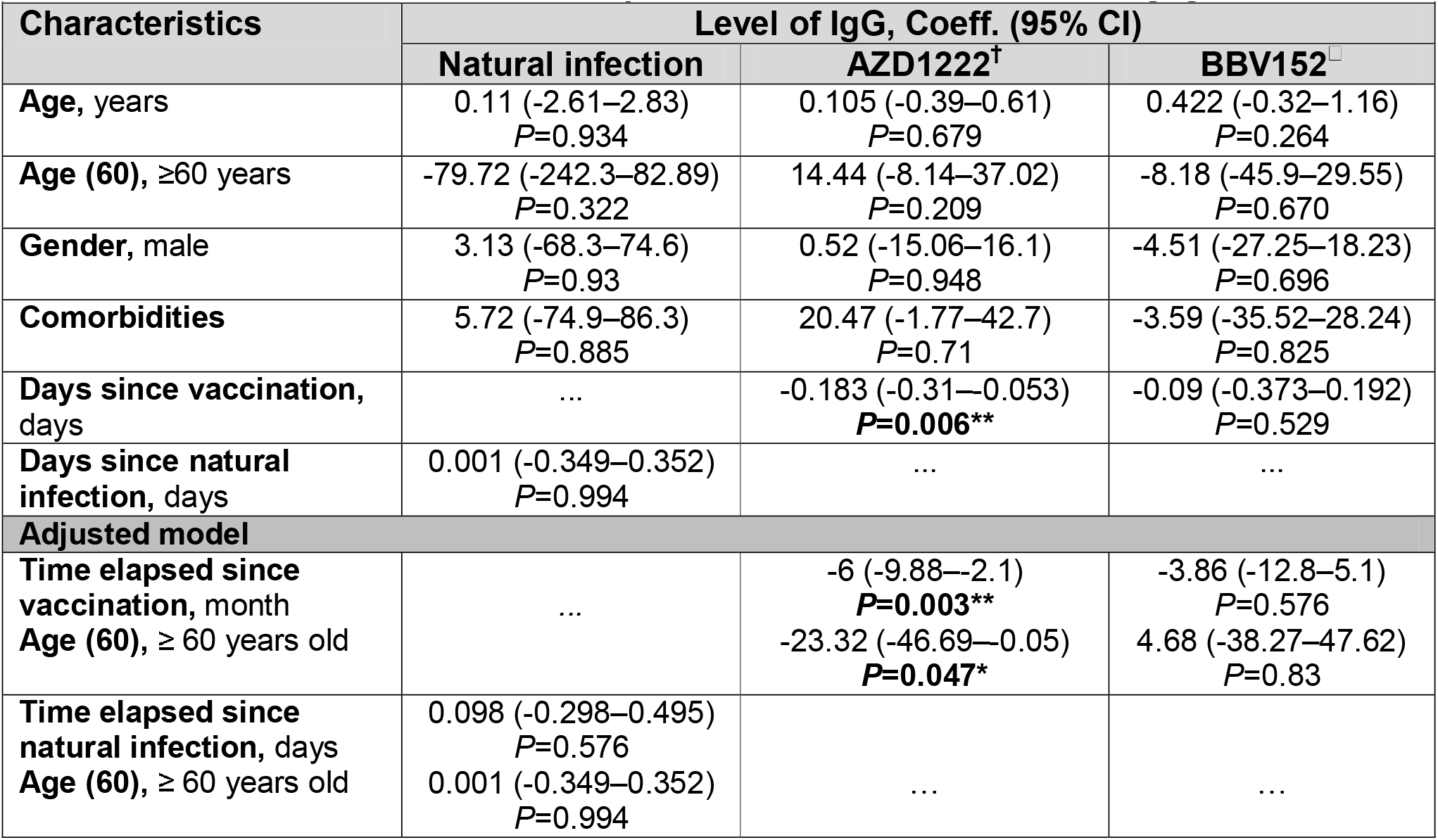
Factors associated with decay of anti-SARS-CoV-2 neutralizing IgG levels. Factors associated with decay of anti-SARS-CoV-2 IgG levels over time among study participants who experienced a natural infection versus vaccination with the commercial AZD1222^†^ and BBV152^□^. Since circulating IgG levels will decay gradually over time following a natural infection and/or vaccination, the levels of the antibodies were analyzed by using a linear regression model controlling for the days since participants become exposed to a natural infection or has completed the second dose of vaccination. Coeff., coefficient; CI, confident interval; month, define as 30 days. *, **, ***, ****, represent *P* value <0.05, <0.01, <0.001, <0.0001 respectively. The Hosmer–Lemeshow value for this model was P=0.634. **Note:** Included only age (60 years) in the adjusted model but not comorbidity. This is because old age and comorbidity are highly associated. ^†^AZD1222 contains a replication-deficient chimpanzee adenovirus, which has a genetic material similar to that of SARS-COV-2. ^□^BBV152 is a whole-virion inactivated vaccine.

**Figure 3:**
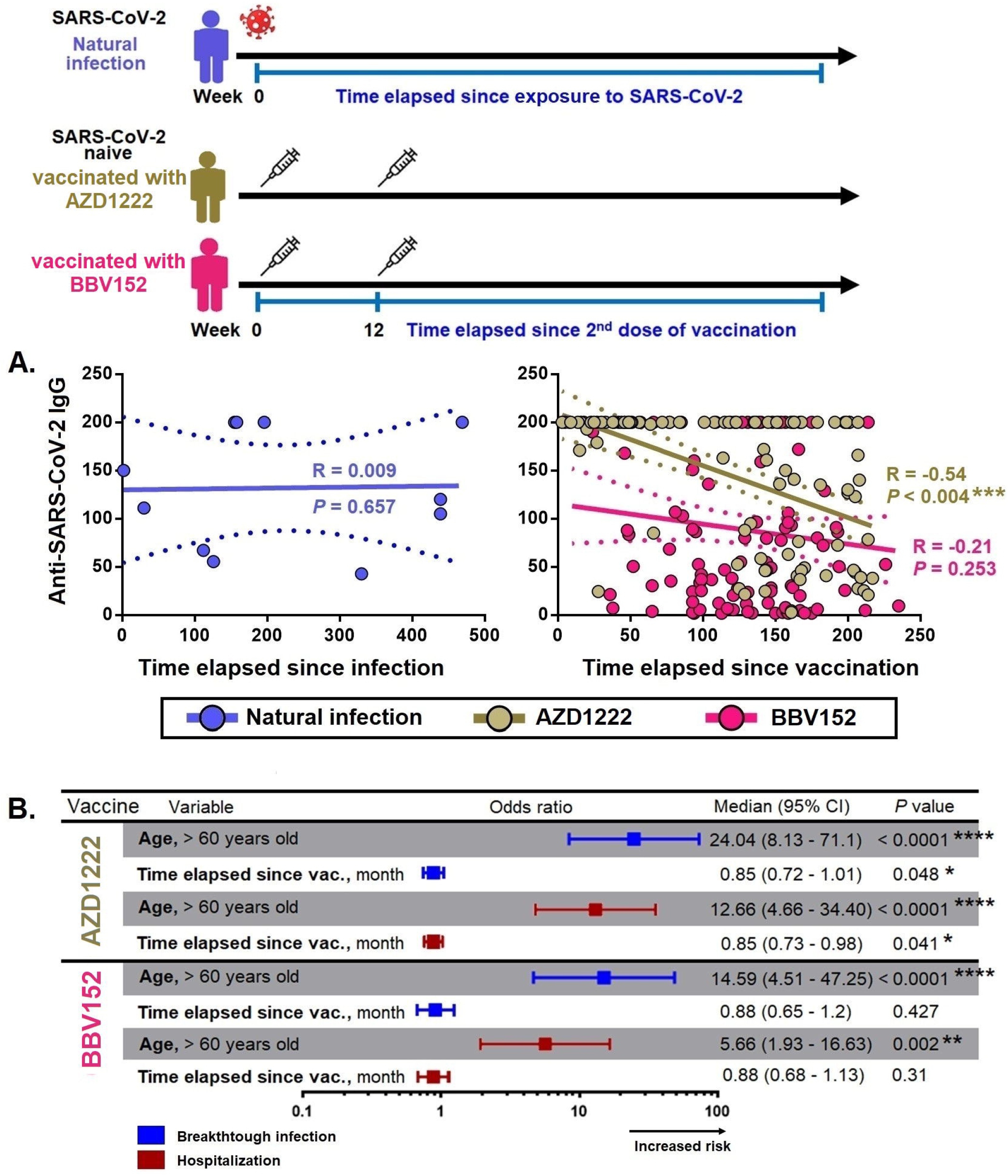
Factors associated with decay of anti-SARS-CoV-2 neutralizing IgG antibodies. **A)** Spearman correlation between the levels of anti-SARS-CoV-2 IgG with the time elapsed since exposure to infection/vaccine administration. **B)** Binary regression models assessing the association between age (>60 years) and time elapsed since vaccination with breakthrough infection and hospitalization. Odds ratios were displayed in a forest plot; median and 95% CI were calculated. CI, confidence interval; month define as 30 days. *, **, ***, *** represent *P*<0.05, <0.01, <0.001, <0.0001 respectively.

## Discussion

In this large population-based real-life study, we studied 519 individuals examined for anti-SARS-CoV-2 neutralizing IgG antibody titers following either vaccination or recovery from documented COVID-19 infection. We investigated the factors associated with development of a breakthrough infection, as well as hospitalization, and correlated them with the dynamics of anti-SARS-CoV-2 IgG titers, as well as factors associated with the decay of the antibodies. We found that age and comorbid conditions were the two factors independently associated with development of a breakthrough infection and hospitalization. A combination of both age (>60 years) and underlying comorbid conditions were associated with increased risk for contracting a breakthrough infection and hospitalization by ∼15 and ∼10 times, respectively. Anti-SARS-CoV-2 IgG decay was only observed among recipients of AZD1222, but not BBV152 and those who recovered from a natural SARS-CoV-2 infection. Due to the decay of anti-SARS-CoV-2 IgG, we also reported that the risk of developing a breakthrough infection and hospitalization gradually increased by 0.85 times with each month.

It is pivotal to understand when and how a breakthrough infection with SARS-CoV-2 occurred in fully vaccinated individuals as it is paramount to determine how long the public health measures needs to be in place and whether or not a community required a booster dose [16]. Immunity against viruses works primarily by inhibiting the infection phenomenon either by humoral (e.g., neutralizing antibodies) or by killing the infected cells via cell-mediated immune responses. While a vaccine works by generating immune memory in the form of memory B cells and T cells that permits a more rapid and intensified immune response against secondary infection; most vaccines are not completely designed to prevent exposure or transmission of an airborne pathogen such as SARS-CoV-2. Hence, acquisition of a breakthrough infection is determined by whether the vaccinated individual at the time of exposure has adequate levels of protective immunity to prevent the establishment of an infection [16]. Many factors are known to influence immune surveillance including the age of the host, the dynamics of antibody responses [17], type/nature of vaccine used [4], interval between the vaccine doses [18, 19], underlying comorbid conditions and other health issues (viz., neoplasms and immunocompromised state) [20].

Several studies on the dynamics of anti-SARS-CoV-2 IgG levels post-vaccination and after recovering from a natural SARS-CoV-2 infection have revealed that the antibody levels induced by vaccines generally undergo rapid decay (over the months). One study reported that individuals who received a AZD1222 vaccine had a substantial decline in antibody levels after six months and that the decline was significantly associated with development of a breakthrough infection [21]. Similarly, although individuals who received the BNT162b2 mRNA vaccine had high antibody titers compared to those who had survived a natural infection, the antibody titers experienced a rapid decay by up to 40% for every subsequent month; whereas the decrease of antibody levels was only <5% per month among convalescing individuals [22]. This is consistent with our observation that individuals administered with AZD1222 experienced a more rapid attrition in IgG antibody levels.

B cells that encounter their cognate antigens during an infection upon activation migrate to the center of the B cell follicle, where they form germinal centers (GCs) [23, 24]. Within the GCs, B cells compete for a limited amount of T cell-derived signals, such as cytokines and CD40 ligand that promote further maturation and differentiation into memory B cells or plasmablasts [25, 26]. Some of these plasmablasts will mature in the secondary lymphoid tissue itself into antibody-secreting plasma cells with short life spans; the other plasmablasts may enter into the circulation together with memory B cells, home to bone marrow and other mucosal tissues, where they mature into long-lived plasma cells or memory B cell that reside in these tissues to secrete antibodies for prolonged periods [23, 27, 28]. Although both infection and vaccination can induce memory B cells and plasmablasts that participate in humoral immune response, due to subtle differences in the nature of antigen stimulation, the memory B cells generated in each case may be different. One study compared the memory B cells induced following inoculation of a BNT162b2 mRNA vaccine and recovery from a natural infection and found that the mRNA vaccine induced robust plasmablast responses as compared to a natural infection that more prone to memory B cells, thereby generating more plasma cells as well as better antigen-binding maturation [29]. Another study compared the immune responses generated by both mRNA-based vaccine and the inactivated whole-virion vaccine and reported that the mRNA-based vaccine induced stronger humoral immune responses than the inactivated whole-virion vaccine [30]. Further, the inactivated whole-virion vaccine induced significantly higher levels of IFN-γ response in CD4^+^ and CD8^+^ T cells as compared to that by the mRNA-based vaccine [30]. Because T cell-derived signals (e.g. cytokines, ligands) are pivotal in promoting B cell maturation in germinal centers [25, 26], the inactivated whole-virion vaccine likely induced long-lived plasma cells and memory B cells.

Of note, there appears to be a fundamental difference between the adenovirus-based vaccine (AZD1222) and the mRNA-based subunit vaccine. Since adenovirus-based vaccine presents as a whole virus, it likely induces B cell maturation much similar to inactivated SARS-CoV-2, and hence be able to generate long-lasting antibody responses. However, because this is an adenovirus-based vaccine and given that adenovirus is common in the population, the presence of anti-adenovirus neutralizing antibodies and anti-adenovirus specific T cell response can prevent the vector from transducing the target cells, thereby limiting the efficacy of the vaccine [31]. In fact, this likely could be a universal concern with all vaccines because of the presence of T and B cross-reactive memory responses to seasonal coronaviruses. Hence, it is difficult for a subunit vaccine that uses spike protein alone without adjuvants to induce long-lived plasma cells [32]. These warrants improved vaccine formulations with suitable adjuvants to enhance antigenic stimulation.

Our study also highlighted that the elderly age group (>60 years) and those with underlying comorbid conditions not only risk the acquisition of a breakthrough infection and hospitalization, but also that individuals administered with the AZD1222 suffered an accelerated decline in anti-SARS-CoV-2 IgG levels. This likely could stem from an ongoing aging phenomenon of the immune system, known as immunosenescence (immune aging), where the generation of new T cells progressively declines due to thymic atrophy. This decline is compensated for by the homeostatic proliferation of mature T cells in the periphery. Eventually, the continually replicating mature T cells become exhausted due to shortening of telomere [33] leading to expression of senescent phenotypes such as loss of co-stimulatory receptors (e.g. CD28, CD69) [34, 35], and de novo expression of inhibitory receptors such as killer-like immunoglobulin receptors (KIR) and PD-1 [33, 35]. Our findings underpin that the elderly with underlying comorbid conditions is a high-risk population that requires more medical attention, and specific measures to boost anti-SARS-CoV-2 immune responses (such as administering a second booster dose of the vaccines) in these groups are urgently warranted.

The limitation of this study is that we not take genetic variant of SARS-CoV-2 [36-39], especially the Delta and Omicron variants that are known to engender high viral loads and high transmissibility into consideration. Notwithstanding, our study provides detailed information in a relatively large cohort of both vaccinated and convalescent individuals recovering from SARS-CoV-2 infection. Our results showed that the declining slope of neutralizing antibodies in AZD1222-vaccinated individuals is much steeper than in convalescent individuals and those who received the BBV152. We also provided estimation of the decline rate as well as corresponding breakthrough infection and hospitalization risks by considering age, underlying comorbid conditions and time-scales into account.

## Data Availability

All data produced in the present study are available upon reasonable request to the authors

## Author Contributons

The study was designed by STS, YKY, GS, KN, SR, YKY, and EMS. STS, GS, and KN provided regulatory oversight. STS and KN provided project management. STS, GS, KN, MR, and SR collected study data and oversaw participant visits. Participant data analysis and interpretation were done by YKY, HYT, YZ, EMS, SR and ML. Patient data collected and analysed by STS, KN, GS, KV, PJ, DR, SP, SR, YKY, HYT, YZ, EMS, SR, and ML and was interpreted by STS, KN, SR, YKY, ML, and EMS. STS, YKY, HYT, KN, GS, and EMS wrote the manuscript. Data were accessed and verified by STS, GS, and SR. All authors contributed to the reviewing and editing of the report and approved the final version.

