## Supplementary Table 1 and 2 for "Factors associated with the decay of anti-SARS-CoV-2 neutralizing antibodies among recipients of an adenoviral vector-based AZD1222 and a whole-virion inactivated (BBV152) vaccine in Chennai, India: a prospective, longitudinal, cohort study"

**Supplementary Data**

| **Supplementary Table 1:** Clinico-demographic characteristics of the study cohort. | | | | | | | |
| --- | --- | --- | --- | --- | --- | --- | --- |
| **Characteristics** | **Total** | **Unvaccinated** | **Vaccinated (n=467)** | |  | ***P* value** | |
|  |  |  | **AZD1222** | **BBV152** |  | a. | b. |
| **Number of participants,** no. (%) | 519 (100) | 52 (10) | 259 (55.5) | 208 (44.5) |  | … | … |
| **Age,** year; median (IQR) | 34 (26–52) | 32 (25.5–42.7) | 35 (27–53) | 34 (25–52) |  | 0.419 |  |
| **Gender,** (male) no, (%) | 248 (47.8) | 27 (51.9) | 115 (44.4) | 106 (51) |  | 0.855 |  |
| **Healthcare workers,** no. (%) | 237 (45.7) | 18 (34.6) | 117 (45.2) | 102 (49) |  | 0.097 |  |
| **Comorbidities,** no. (%) | 74 (14.3) | 7 (13.5) | 36 (13.9) | 31 (14.9) |  | 0.775 |  |
| *Hypertension,* no. (%) | 37 (7.1) | 4 | 19 | 14 |  | … |  |
| *Diabetes,* no. (%) | 24 (4.6) | 2 | 10 | 12 |  | … |  |
| *Heart Disease,* no. (%) | 4 (0.8) | 0 | 3 | 1 |  | … |  |
| *Others*^†^*,* no. (%) | 9 (1.7) | 1 | 4 | 4 |  | … |  |
| **SARS-CoV-2 infection;** no. (%) | 176 (33.9) | 27 (51.9) | 82 (31.7) | 67 (32) |  | 0.016^a^* | 0.25 |

**Note:** *Others^†^* consist of asthma=4, arthritis=2, cancer/cured cancer=2, chronic lung disease=1. a) Comparison between three groups, i.e., Unvaccinated, AZD1222 and BBV152; b) Comparison between, AZD1222 and BBV152. * *P*<0.05.

| **Supplementary Table 2:** Clinical and demographic characteristics associated with hospitalization. | | | | | | | |
| --- | --- | --- | --- | --- | --- | --- | --- |
| **Characteristics** | **Univariate model** | | | **Multivariate model** | | | |
|  | **Coeff.** | **(95% CI)** | ***P* value** | | **Coeff.** | **(95% CI)** | ***P* value** |
| **Age** (5 years) | 1.312 | (1.224 – 1.401) | <0.0001 **** | | 1.267 | (1.179 – 1.362) | <0.0001 **** |
| **Gender** | 1.451 | (0.988 – 2.128) | 0.058 | | 0.771 | (0.509 – 1.168) | 0.219 |
| **Vaccine status** | 0.653 | (0.326–1.31) | 0.229 | | … | … | … |
| **Vaccine type**  AZD1222  BBV152 | 0.687  1.095 | (0.331 – 1.425)  (0.732 – 1.637) | 0.313  0.659 | | …  … | …  … | …  … |
| **Comorbidity** | 3.973 | (2.392 – 6.601) | <0.0001 **** | | 1.930 | (1.098 – 3.394) | 0.022 * |

**Note:** Coeff., coefficient; CI, confident interval. *, **, ***, ****, represent *P*<0.05, <0.01, <0.001, <0.0001 respectively.
